# Spatial distribution of *Culex* mosquitoes across England and Wales, July 2023

**DOI:** 10.1101/2025.02.28.25322982

**Authors:** Emma Widlake, Roksana Wilson, Jack Pilgrim, Alexander G. C Vaux, Jola Tanianis-Hughes, A. Haziqah-Rashid, Ken Sherlock, Agata Delnicka, Amelia Simpson, Anthony J. Abbott, Colin J. Johnston, Jude Martin, Kendall Barlow, Eloise Aliski, Saffron Shiels, Sara Gandy, Sarah M. Biddlecombe, Luigi Sedda, Jolyon M. Medlock, Matthew Baylis, Marcus S. C. Blagrove

## Abstract

**Background:** With medically important arboviruses such as West Nile virus (WNV) circulating in Europe and Usutu virus (USUV) currently present in the UK, it is imperative to identify areas in the UK at risk of establishment and spread of these viruses. Here, we describe a comprehensive nationwide field surveillance study conducted during July 2023 to map the distribution of the WNV and USUV competent vectors: *Culex pipiens* biotype *pipiens*, *Culex pipiens* biotype *molestus*, and *Culex torrentium*, across England and Wales.

**Methods:** Mosquitoes were sampled for 3 trap nights (3TN) at two hundred sites in rural, urban and suburban settings, selected using a lattice plus close pairs surveillance design. Field caught samples were analysed using morphological and molecular approaches.

**Results:** A total of 2,157 adult mosquitoes of the *Culex* genus were collected. *Culex pipiens* biotype *pipiens* dominated the resident *Culex* populations, comprising 1,478 (95.8%) out of 1,543 mosquitoes with confirmed molecular species identity. *Culex torrentium* were present in much lower numbers, with only 38 (2.5%) identified. Only five of the biotype *molestus* (0.3%) were identified in this study, however these were found in localities outside of London and in a rural location, with the biotype previously having been associated with human-made habitats. This study also found that hybrids of the pipiens and molestus biotypes are more widespread than previously recorded. In total, 22 hybrids (1.4%) were identified from counties as far west as Cornwall and as far north as Suffolk.

**Conclusions:** Coupled with reviews of previous UK *Culex* sp. surveys, this study provides essential data for risk modelling of WNV and USUV, furthering the UK’s preparedness for incursions of vector-borne diseases in the future.

## Background

Arthropod-borne viruses (arboviruses) of human and animal health significance are a current and growing threat in Europe. Warming climates are leading to the expansion of the suitable range of both arbovirus vectors, and the viruses themselves and likely impacting the spread, density and seasonality of the vectors [1]. West Nile virus (WNV), a medically important arbovirus, has spread throughout Europe in the last few decades. From the beginning of 2024 to September 2024, 16 countries in Europe have reported human cases of WNV infection, including France, Germany, Italy, Romania and Greece [2]. Whilst WNV has not yet been detected in the UK, models predict local transmission in the latter half of the century [3]. Usutu virus (USUV) was first isolated in resident dead birds and *Culex pipiens s.l* mosquitoes in London in 2020, with further detections suggesting it may have become established [4–6]. Both viruses are co-circulating in at least twelve European countries [7]. With increasing frequency of outbreaks in Europe, and the presence of these viruses in the UK or neighbouring countries such as France and the Netherlands, there is a growing risk to people in the UK.

WNV and USUV are *Orthoflaviviruses* grouped in the Japanese Encephalitis Complex. USUV is maintained through an enzootic cycle between mosquitoes and avian hosts. Blackbirds (*Turdus merula*) are highly susceptible to USUV and fatalities have been reported with a decrease in reported sightings in the UK in recent years coinciding with the discovery of the virus [5,8]. Similarly, WNV is well known for infecting wild birds and in 2023 eight European Union countries reported 251 outbreaks of WNV among birds [9]. WNV and USUV can both infect a range of mammals, including humans, following infected mosquito bites [10–14]. These mammalian hosts are not part of the transmission cycle and as such are viewed as “dead end” hosts (i.e. the level of viraemia is insufficient to infect blood-feeding mosquitoes). With both viruses infecting avian species, migratory birds can introduce the viruses to new areas independent of the dispersal of infected mosquitoes, although there is also a small chance of infected mosquitoes being introduced to new areas via aircraft [15,16].

Whilst 70-80% of WNV cases in humans are asymptomatic, WNV can cause febrile disease with fever and flu-like symptoms [17]. A small proportion of patients develop WNV neuroinvasive disease characterised by acute flaccid paralysis and meningoencephalitis [18]. Human infection with USUV is rare [19]. Based on current data, USUV generally produces a mild or asymptomatic infection, however, it can lead to neuroinvasive disease characterised by meningitis, encephalitis or meningoencephalitis in both healthy and immunocompromised patients [20]. Globally, only two deaths attributed to USUV (in individuals who are immunocompromised or have co-morbidities) have been recorded [21,22].

The *Culex* genus includes competent vectors for WNV and USUV [23–28]. UK *Culex* species include: *Culex territans*, *Culex modestus, Culex torrentium* and *Culex pipiens s.l* (hereafter *Cx. pipiens* complex) a mosquito complex which consists of *Cx. pipiens* biotype *pipiens* (hereafter *Cx. pipiens*), *Cx. pipiens* biotype *molestus* (hereafter *Cx. molestus*) and hybrids of *Cx. pipiens* and *Cx. molestus* (hereafter *Cx. pipiens/molestus* hybrids) [29,30]. *Cx. torrentium* and the *Cx. pipiens* complex can only be reliably differentiated using molecular techniques [31] which are rarely performed in surveys. Consequently, data on the occurrence of *Cx. pipiens* complex species, biotypes and hybrids are scarce [32].

In the UK, the two *Cx. pipiens* biotypes show differing characteristics that may allow them to contribute separately to the transmission and maintenance of WNV and USUV in human and avian populations*. Cx. pipiens* is widespread and abundant, surface-dwelling and enters diapause during the winter months. It commonly breeds in a range of container and natural aquatic habitats. Females require a blood meal to lay eggs (anautogenous), are primarily ornithophilic [29,30,33–34] and infrequently feed on humans [35,36]. Based on their feeding preferences, *Cx. pipiens* is likely to be an enzootic vector of WNV and USUV in the UK. On the other hand, *Cx. molestus* is thought to be less common and more focal in its distribution, primarily subterranean (e.g. mainly limited to underground railway tunnels, mines and basements) and remains active during the winter [29,24,37]. Above-ground *Cx. molestus* have previously been reported in Mogden (sewage works) and Beckton (houses) in London, at a farm location in east London [38] and in Neston, Cheshire [33,38,28]. Females do not require a blood meal to lay their first batch of eggs (autogenous) and female mosquitoes prefer humans as hosts (anthropophagic) [29,34,37]. *Cx. molestus* in mainland Europe are reported to be opportunists that feed on both birds and humans [39, 40, 41]. *Cx. molestus* in the UK have not yet been recorded feeding on birds, although the species is under-studied.

In southern Europe, both biotypes can be found in the same habitat, and they occasionally hybridise [39, 40,41]. In the UK, however, there are assumed to be few opportunities for hybridisation because the two biotypes are usually found in separate above- and below-ground habitats, although *Cx. molestus* adults have been occasionally recorded biting at the surface [42]. Hybridisation may affect the risk of WNV or USUV transmission, because hybrids are potentially more opportunistic in their feeding than either biotype and therefore could bridge the viruses between birds and humans [43, 32]. The remaining UK *Culex* species: *Cx. territans, Cx. modestus* and *Cx. torrentium* are less common or even rare. *Cx. territans* primarily feeds on amphibians and infrequently bites humans and birds [44]. Its feeding preferences and scarcity means it is unlikely to substantially contribute to WNV and USUV transmission [34]. *Cx modestus* is mostly limited to wetlands in north Kent and the coast of Essex, however, it is highly abundant at these sites [45], although it has recently been confirmed at a number of wetland sites in Cambridgeshire and on the Hampshire/Sussex coast (UKHSA, in prep). The species routinely feeds on both humans and birds, making it a potential bridge vector of WNV and USUV to humans [36,46]. *Culex torrentium* is understudied and much of the information on this mosquito is anecdotal [47]. It is considered an ornithophagic species [48]. Based on their feeding preferences, *Cx. torrentium* is likely to be an enzootic vector of WNV and USUV. Wetland and marsh areas have historically been prominent areas for WNV outbreaks to begin due to the concomitance of high numbers of mosquitoes, migratory birds and often proximity to human and equine populations [49,50].

Previous outbreaks of WNV in Europe have shown high numbers of cases in areas where the virus has previously been detected, potentially due to overwintering and local transmission of the virus [51,52]. This suggests that following establishment, efforts to prevent recurrent transmission will be difficult. The importance of understanding areas most at risk of WNV and USUV transmission cannot be understated, and the role of mosquito surveillance in this effort is essential, particularly in determining the relative abundance of potential enzootic vectors such as *Cx. pipiens* and *Cx. torrentium* to sustain bird to bird transmission, but also in determining the distribution and abundance of potential bridge vector *Culex* species, such as *Cx. molestus,* its hybrids with *Cx. pipiens*, and *Cx. modestus* [45]. Identifying regional variation in the distribution of mosquitoes capable of transmitting these viruses allows us to understand where these viruses may become established and devise targeted preventative measures and vector control strategies. Whilst *Culex* mosquitoes have been studied extensively in other parts of the world, comprehensive mapping of their distribution and species diversity and abundance across England and Wales remains limited, particularly for the *Cx. pipiens* complex as they cannot reliably be separated morphologically.

This study aimed to conduct a comprehensive analysis of *Culex* species diversity, and presence and absence, across England and Wales during peak *Culex* season in July 2023. Adult and immature mosquitoes were collected from a total of 200 sites selected using a lattice with close pairs spatial design, and members of the *Culex* genus were identified using a combined morphological and molecular approach that could differentiate *Cx. torrentium*, *Cx. pipiens*, *Cx. molestus* and *Cx. pipiens/molestus* hybrids. Regional differences in the occurrence of these mosquitoes were determined and results were compared with previous *Culex* surveys in the UK. The study’s findings will better inform risk assessments on USUV and WNV in relation to the potential for enzootic and bridge vector transmission as well as highlight future research questions on the ecology of *Cx. molestus* and *Cx. pipiens*/*molestus* hybrids.

## Methods

### Field collection

Sampling point locations across England and Wales were chosen using a “lattice plus close-pairs” design that combines points spread across a regular lattice and random points as close pairs. This design is efficient for parameter estimation of geostatistical models [53,54]. To be representative of the UK Centre for Ecology and Hydrology’s (UKCEH) 21 land cover classes (Figure 1), the distribution of the 200 sampling points met two conditions: (i) 188 of the sampling points were in a lattice while 12 were distributed randomly but close to the lattice sampling points (at a distance of 5 to 10 km); (ii) each land cover contained a number of sampling points proportional to stratum size [55] and not fewer than five (to have a minimum statistical power). The lattice comprised 60 cells that were mostly 50 by 50 km and sites were positioned as close as possible to a regular distance of approximately 50 km. Due to the irregular shape of England and Wales, 6 squares were smaller than 50 x 50km and 12 were larger. Within each square, sampling locations were defined by squares of 2 x 2 km, either at the centre of the land cover class or at least 1 km apart from other land classes to reduce cross-over.

**Fig. 1.**
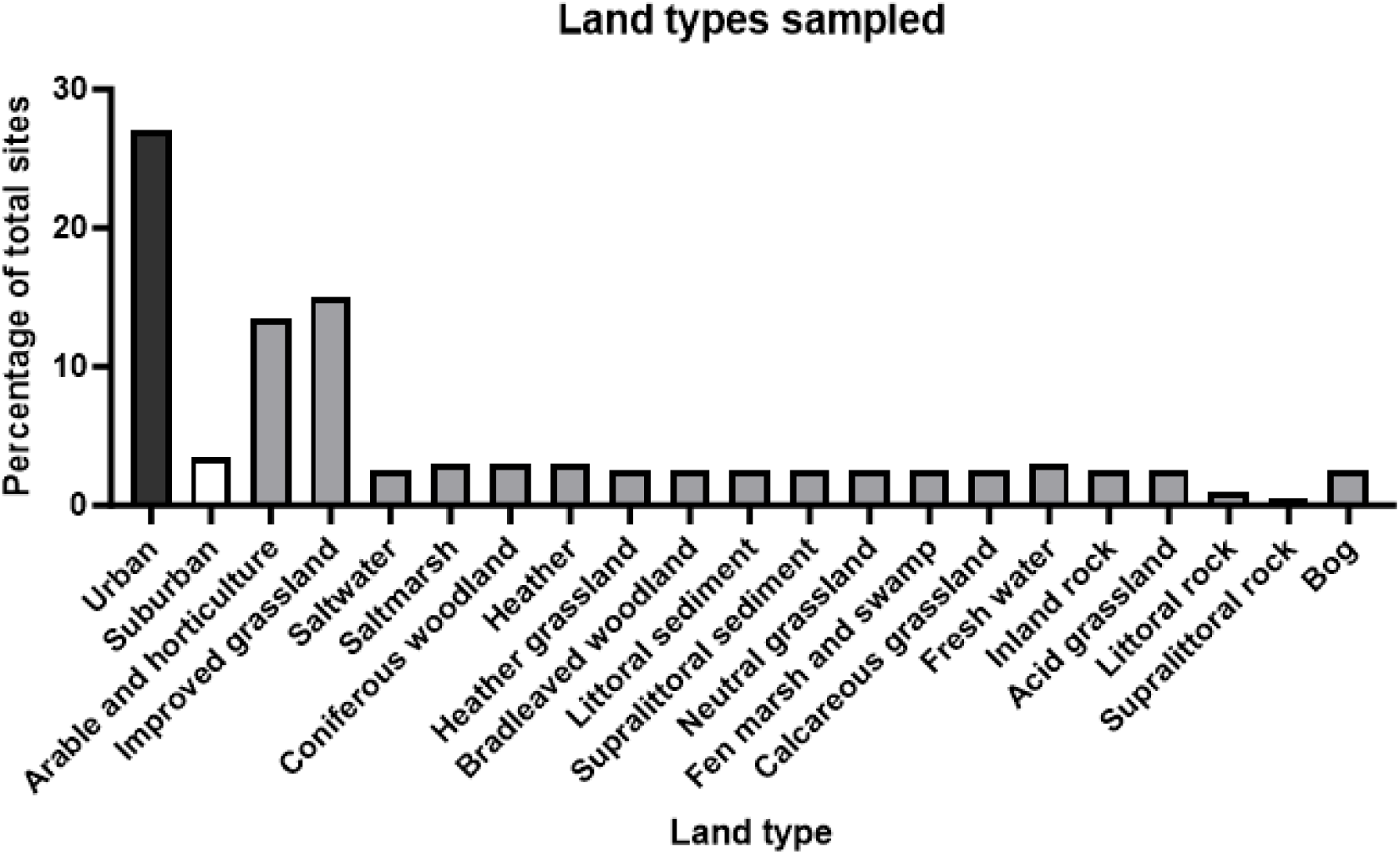
The percentage of each land type (n = 21) visited across the total of 200 sites. These land types are designated to a site based on the majority land coverage within a 1×1 km square in which the trap site was selected. The land coverage was defined using the UKCEH land cover class [56]. For the rest of the manuscript, results have largely been divided into rural (any land type not including urban and suburban), urban and suburban.

The survey was conducted by two teams from the University of Liverpool covering the northern parts of England and Wales (100 trap sites) and three teams from the UK Health Security Agency (UKHSA) covering sampling points in the southern parts of England and Wales (100 trap sites). At each previously determined 2 x 2 km site, the most appropriate location for *Culex* collection was identified based on shelter and proximity to water, if available. At each designated site, BG-PRO® adult mosquito trap (Biogents AG, Regensburg, Germany) with BG-Lure® (Biogents AG) and BG-CO_2_ Generator®, and a BG-GAT® (Gravid Aedes Trap; Biogents AG) were deployed and operated for a 72-hour period (three-days). An average of 12.5 (±SE 2.2) traps were set per day. Upon collection, mosquito specimens were preserved at 4°C until transfer after no longer than 72 hours to −80°C for storage. Mosquitoes of the *Cx. pipiens* complex, and *Cx. modestus,* were identified morphologically [37], while molecular techniques were used to identify *Cx. pipiens*, *Cx. molestus*, *Cx. pipiens/molestus* hybrids and *Cx. torrentium*.

### Traps

BG-PRO traps were assembled according to the manufacturer’s instructions, utilising the Sentinel style method. BG-CO₂ Generators were set according to manufacturer instructions and placed into the insulated carry bag next to the trap. Biogents BG-CO₂ powder contains various yeast strains and nutrients for production of CO₂ over a 24-hour period, with highest output in the first 14 hours. A BG-Lure was also added to the BG-PRO to further attract mosquito species. A battery pack tested to last at least 72 hours was attached to the BG-PRO to keep the fan in motion (Energizer UE30058 30,000mAh Power Bank, TennRich International Corp, Taoyuan). These traps were run for 72 hours.

The BG-GAT trap setup involved the preparation of hay infusion water two weeks prior to fieldwork to attract gravid females for egg-laying. Hay infusion was made as described by [58] (200L water, 9kg dried hay, 10g lactalbumin and 10g brewer’s yeast (first described by [57]) and this was mixed 1:1 with water for the trap. These were left for 72 hours. Normally, a thin mesh is placed over the water, however this was removed to allow mosquitoes to lay eggs in the traps, to enable identification of mosquito eggs. A small sheet of sticky paper was taped to the opening of the trap to catch mosquitoes entering or exiting. Both the BG-PRO and BG-GAT were placed within 1 m of each other (Figure 2).

**Fig. 2.**
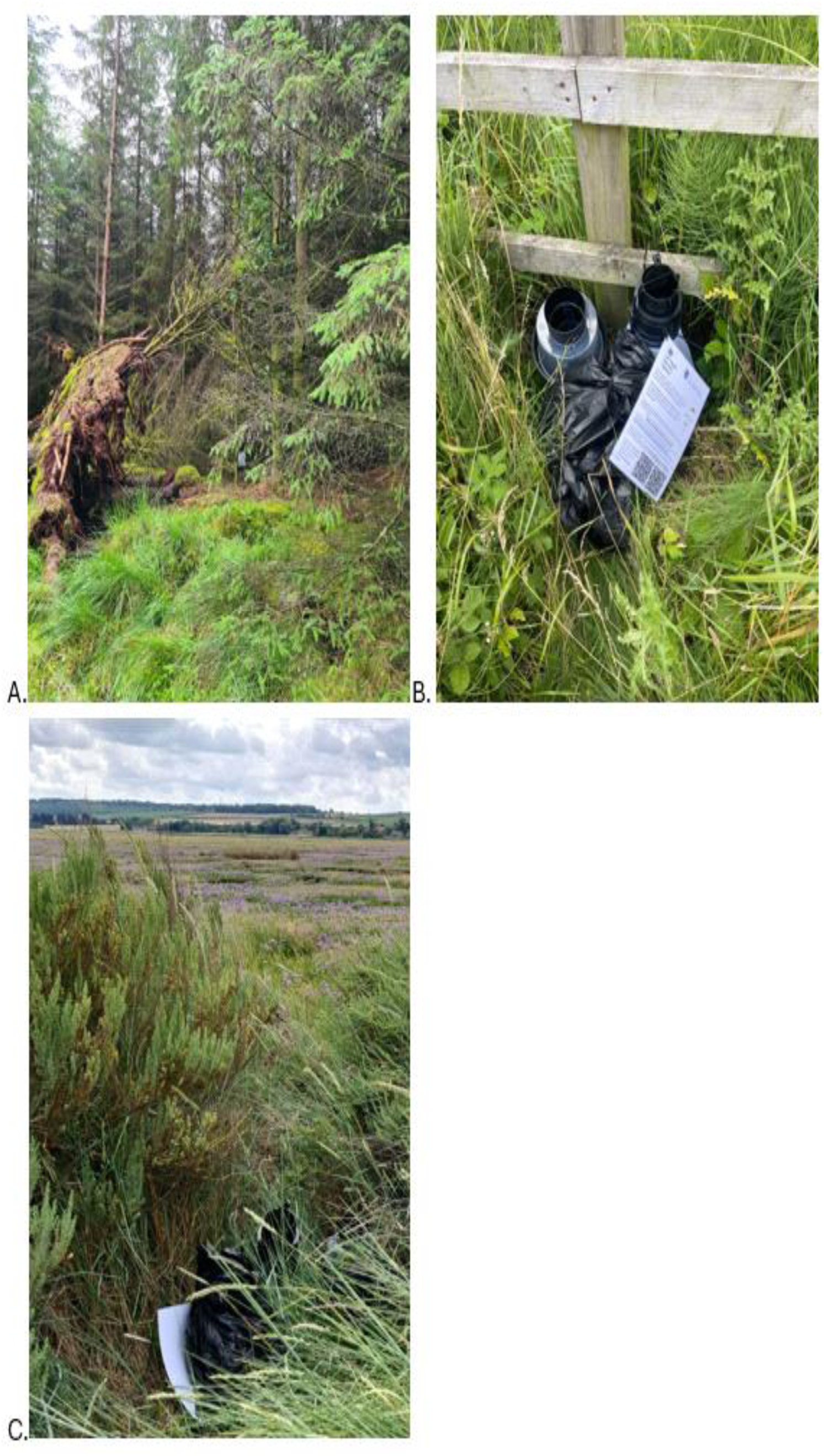
Example photos of the BG-PRO and BG-GAT traps set up during the July 2023 *Culex* field collections. At all sites, both traps were placed within 1 m of each other. The traps were normally placed in sheltered areas. A. view of trap site from public footpath (the trap is visible just below centre), B and C. close up images of traps placed next to one another.

### Larval and pupal collections

At each trap site, a search for water bodies was conducted. Suitable water bodies were those with the potential to retain water, including water troughs/butts, buckets, tyres and ponds and excluded very temporary water such as rainwater puddles. Larvae were collected from sites where observed and species ID was performed as described below for the adults. First instar larvae from each site were pooled prior to the DNA extraction, whilst all other instars and pupae were processed individually. All larval data presented in this paper was collected from the southern parts of England and Wales.

### DNA extraction

Only female mosquitoes were analysed due to their ability to blood feed and become infected with arboviruses. Mosquito samples were individually bead beaten with 5 mm steel beads at 225 rpm for 2.5 minutes in 90 μL of proteinase K buffer. 10 μL of proteinase K was added to each sample followed by 90 minutes incubation at 60 °C degrees. Samples were stored at −20 °C until DNA extraction the next day.

Samples were subject to DNA extraction using the KingFisher Flex system (Thermo Scientific). Plates were prepared for the KingFisher Flex as per the manufacturer’s instructions. All solutions were from the MagMax CORE kit unless stated otherwise. First, 500 μL of wash solution one was added to each well of a deep well plate. Next, 500 μL of wash solution 2 was added to a deep well plate. After that, 90 μL of elution buffer was added to a standard 96 well plate. A tip comb was added to a standard 96 well plate. Finally, 350 μL of lysis solution, 350 μL of binding solution, 20 μL of magnetic beads and 100 μl of sample were added to a deep well plate, with one well per sample. Kingfisher Flex extraction protocol uses the MagMAX_CORE_Flex_no_heat protocol. DNA samples were stored at 4 °C until initial PCRs were completed.

### Species identification polymerase chain reaction

PCR reactions were set up using the Thermofisher Dreamtaq kit as per the manufacturer’s instructions to a total of 13 μL: 6.25 μL DreamTaq master mix, 1.25 μL forward primer, 1.25 μL reverse primer, 3.25 μL H2O and 1 μL DNA.

For *Culex pipiens* complex vs *Cx. torrentium*, primers targeting the COI gene were used, forward primer: 5’-CAACATTTATTTTGATTTTTTGG-3’ and reverse primer: 5’-TCCAATGCACTAATCTGCCATATTA-3’. Bands for both species are produced at approximately 830 bp and further restricted for differentiation (see digests below). The PCR protocol was as per [58].

To differentiate between *Cx. pipiens*, *Cx. molestus* and *Cx. pipiens/molestus* hybrids, the following primers were used: forward primer: 5’-GATCCTAGCAAGCGAGAAC-3’, reverse primer 1: 5’-CATGTTGAGCTTCGGTGAA-3’ and reverse primer 2: 5’-CCCTCCAGTAAGGTATCAAC-3’. *Cx. pipiens* returned a band of approximately 200 bp, *Cx. molestus* a band of 250 bp and *Cx. pipiens/molestus* hybrids two bands of 200 and 250 bp in length. The PCR protocol was as per [59].

### Restriction Digestion

To differentiate between *Cx. pipiens* complex and *Cx. torrentium*, PCR products were digested with FspBI and SspI individually as follows: 8.5 μL H2O, 1 μL fast digest buffer, 0.5 μL enzyme and 5 μL PCR product. FspBI reactions were incubated at 37 °C for 10 minutes before heat inactivation at 80 °C for 5 minutes. SspI reactions were incubated at 37 °C for 10 minutes before heat inactivation at 65 °C for 5 minutes. Digest products were run on a 2 % agarose gel. In the first digest, FspBI cuts *Cx. torrentium* PCR product into two bands of 620 and 210 bp in length whilst any *Cx. pipiens* complex bands will remain uncut. In the second digest, SspI cuts *Cx. pipiens* complex into two bands of 620 and 210 bp in length whilst *Cx. torrentium* bands remain uncut. Use of FspBI alone is enough for identification, but use of both enzymes is recommended [58].

### Mosquito association to land cover types

To assess tendencies for *Culex* species to be found in specific land cover types, we first reduced the 21 land cover types to three categories (rural, urban, suburban). We then compared the number of sites that were positive and negative for each *Culex* species, and the total number of each species caught, in the three categories, using the Fisher Exact Test. This test was used instead of Chi-square as some expected cell frequencies were less than 5.

### Mosquito association to human populations and artificial structures

Due to the small sample size of *Cx. molestus*, the association between *Cx. pipiens* and *Cx. molestus* with human density and artificial structures was estimated by employing a Monte Carlo test [60] for species-environment correlations. In practice, this is an ecological case-control comparison where for each iteration of the Monte Carlo test a sub-sample of the same size of *Cx. pipiens* and *Cx. molestus* trap counts and their absences are randomly selected and their correlation with human densities and artificial structures are calculated. With this approach, the sample size for cases (mosquito counts) and controls (mosquito absence) are kept identical through each iteration. The Monte Carlo test was run 999 times and the mean association (from the full set of correlations), including 95% confidence intervals, was produced [60]. Buildings and other artificial structures (parking, power structures, facilities etc) were extracted from OpenStreetMap [61], while human population density (at 30 m spatial resolution) was obtained from Facebook/Meta [62]. Distances between any trapping location point and the centre of artificial structures were calculated using Euclidean distance. Densities of structures and human population were estimated within a 500 m buffer around each trapping point location.

### Probability maps

Models for confirmed *Cx. pipiens* and *Cx. torrentium* (*Y_s_*) at a location, (*s)*, were fitted by employing a Poisson generalised linear mixed model:

*Y*_*s*_ = Poisson(*μ*_*s*_)
*log*(*μ*_*s*_) = *α*_*u*_ + *β***X**_*s*_ + *ε*_*s*_
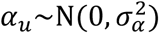
*ε*_*s*_∼MVNormal(0, **Σ**_*ε*_)

**Table 1:** Covariates used in the study. For each covariate, monthly mean, minimum, maximum and standard deviation were calculated (https://cds.climate.copernicus.eu/cdsapp#!/dataset/reanalysis-era5-land?tab=overview).

| Variable | Code | Unit | Source | Resolution |
| --- | --- | --- | --- | --- |
| Where $\alpha$ is a group-level random effect normally distributed with mean equals to 0 and variance $\sigma^2$, with group $u$ defined as either by urban or rural, $\beta$ is a vector of coefficients for each explanatory variable contained in the matrix $\mathbf{X}$ (all the variables reported in Table 1), $\varepsilon$ is the spatial random Gaussian field with covariance matrix $\Sigma_\varepsilon$ constrained by a Matern covariance function. This model was implemented using the sdmTMB package in R software [63]. This modelling approach enables mapping the detection probabilities of *Cx. pipiens* and *Cx. torrentium* [64]. | | | | |
| Horizontal speed of wind towards the east, at a height of ten metres above the Earth surface. | u10 | metres per second | ERA5-Land hourly data from 1950 to present data, Copernicus's Climate Data Store, by European Centre for Medium-Range Weather Forecasts | Temporal: hourly<br>Spatial: 1100 metres |
| Horizontal speed of wind towards the north, at a height of ten metres above the Earth surface. | v10 | metres per second |  |  |
| Leaf area index, high vegetation (evergreen trees, deciduous trees, mixed forest/woodland, and interrupted forest). | lai_hv | square metre per square metre |  |  |
| Leaf area index, low vegetation (crops and mixed farming, irrigated crops, short grass, tall grass, tundra, semidesert, bogs and marshes, evergreen shrubs, | lai_lv | square metre per square metre |  |  |
| deciduous shrubs, and water and land mixtures). |  |  |  |  |
| Skin (Earth surface) temperature | skt | kelvin |  |  |
| Potential evapotranspiration | pev | m |  |  |
| Total precipitation | tp | metre |  |  |
| Volume of water in soil layer 1 (0-7cm) | swvl1 | cubic metre per cubic metre |  |  |
| Gridded population density | pop_density | number of people per square kilometer | Data for Good at Meta (from the HDX) | 2019<br>Spatial: 30 metres |
| Land cover | land_cover | --- | CEH | Spatial: 25 metres |
| Elevation | elevation | metre | U.S. Geological Survey | Spatial: 450 metres |
| Buildings | Number of buildings 500m around the sampling location.<br>Mean distance from buildings.<br>Minimum distance from buildings. | binary (presence/absence) | Openstreetmap/OSM |  |

## Results

### Spatial distribution of *Culex* mosquitoes across England and Wales in July 2023

All mosquitoes were caught in the BG-PRO traps.

Spatial analysis revealed several key findings regarding the distribution and density of *Culex* mosquitoes across different regions and habitats in England and Wales. From a total of 100 northern and 100 southern sites, 78 (39%) sites had no mosquitoes, 5 (2.5%) had non-*Culex* mosquitoes only (*Aedes cantans/annulipes*, *Aedes cinereus*, *Coquillettidia richiardii, Culiseta litorea* and *Culiseta morsitans*), and 117 (58.5%) had *Culex* mosquitoes (alone or with other mosquito species). A list of all mosquito species collected is shown in Supplementary Table S1. Of the 117 *Culex* positive sites, 68 (58.11%) were found in the south and 49 (42.24%) in the north. There was a greater likelihood of finding sites positive for *Culex* in the south (Chi-square, *P*<0.01). A total of 2,157 female mosquitoes of the *Culex* genus were collected over three trapping nights (3TN), comprising 1633 collected in the south and 524 collected in the north, with an average of 24.35 and 10.69 per 3TN respectively. This suggests a higher density of *Culex* mosquitoes towards the south of England and Wales (see Figure 3) than the north. The highest density per 3TN was 312 *Culex* mosquitoes, caught in Devon in the southwest of England.

**Fig. 3.**
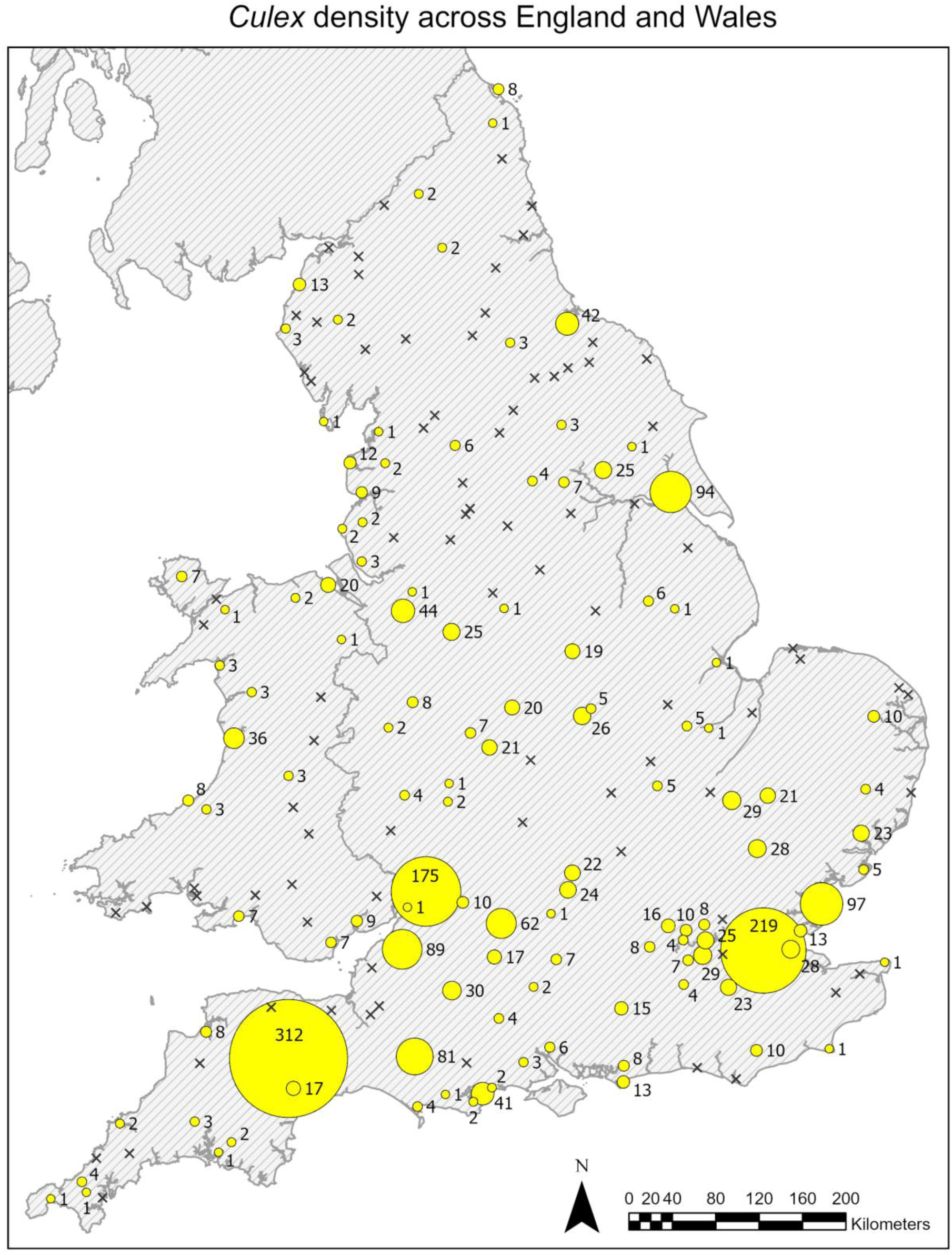
A map showing the density of *Culex* mosquitoes caught across England and Wales in July 2023. Yellow circles represent the trap sites, whilst the size of each circle represents the number of mosquitoes caught at each site per 3TN. The number of mosquitoes caught is noted next to each site. X’s represent sites where no *Culex* mosquitoes were caught. For this map, *Culex* refers to the *Cx. pipiens* complex and *Cx. torrentium*.

A total of 1,543 female mosquitoes were successfully identified to species; 79 were narrowed down to the *Cx. pipiens* complex but did not differentiate the *pipiens* or *molestus* biotypes or their hybrids; and 532 gave ambiguous results or no bands across the three assays. Ambiguous results were samples that gave positive results for two different *Culex* species (i.e. *Cx. pipiens* complex and *Cx. torrentium*). Of the 117 *Culex* positive sites, 15 had samples that could only be confirmed as *Culex* morphologically, giving only one or no results by the species identification (ID) PCR assays. Of the 1,543 samples identified to species, 95.8% were *Cx. pipiens* (n=1,478), 2.5% were *Cx. torrentium* (n=38), 0.3% were *Cx. molestus* (n=5) and 1.4% were *Cx. pipiens/molestus* hybrids (n=22) (see Figure 4).

**Fig. 4.**
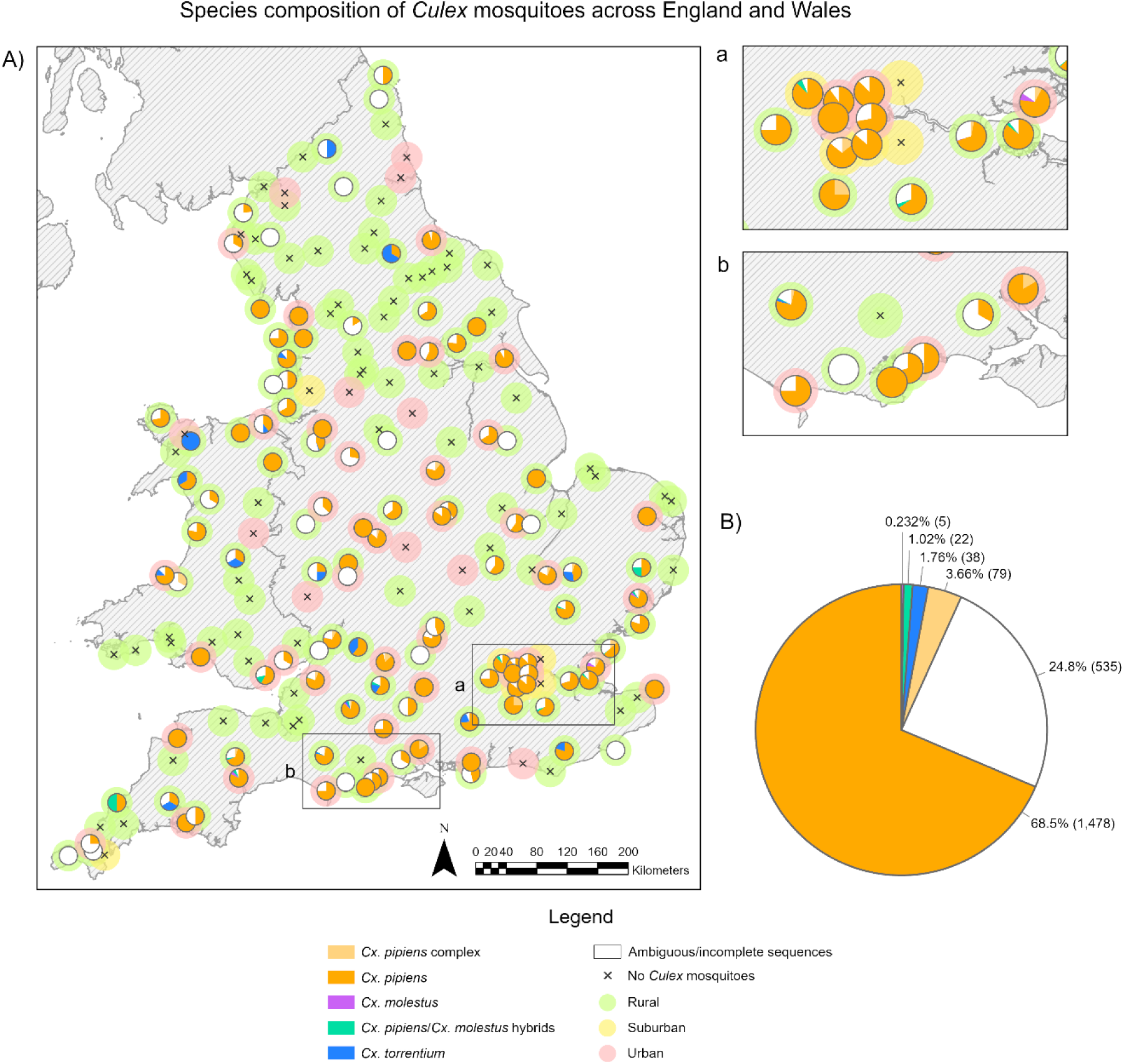
A: A map showing the *Culex* mosquito species composition caught per 3TN and the land cover type of each of the trap sites (rural, urban or suburban). (a) and (b) are zoomed in sections of areas indicated on the map. X’s represent sites where no *Culex* mosquitoes were caught. B: A pie chart showing the species composition of all the *Culex* mosquitoes caught per 3TN across England and Wales in July 2023, with the percentage of each species indicated to the right of each segment. *Cx. pipiens* complex refers to samples identified within the complex but produced no further results to differentiate between *Cx. pipiens* or *Cx. molestus*. Ambiguous results gave results for multiple assays, i.e. positive for both *Cx. torrentium* and *Cx. pipiens*.

### Distribution across rural, urban and suburban areas

Of the 200 sites, 139 (69.5%) were classified as rural, 54 (27%) were urban and 7 (3.5%) were suburban. Of the 1,478 mosquitoes confirmed as *Cx. pipiens*, 910 (61.6%), 524 (35.5%) and 44 (3%) were from rural, urban and suburban locations respectively. Of five individual samples confirmed as *Cx. molestus,* 2 (40%) were caught in rural sites and 3 (60%) were caught in urban sites. Of the 22 mosquitoes identified as *Cx. pipiens/molestus* hybrids, 17 (77.3%), 4 (18.2%) and 1 (4.5%) were from rural, urban and suburban locations respectively. Finally, of the 38 *Cx. torrentium,* 35 (92.1%) were found in rural sites and 3 (5.3%) in urban sites.

Urban sites were significantly more likely to be positive for *Cx. pipiens* (Fisher Exact Test, *P*< 0.01). Nearly 78% of urban sites were positive, compared to only 38.8-42.9% of rural and suburban sites (Table 2). By contrast, there were no significant associations for *Cx. molestus*, hybrids or *Cx. torrentium*, although the latter had a higher frequency in rural sites (12.9%) compared to 3.7 - 0% urban/suburban sites (*P =* 0.13).

**Table 2.** Table showing the number of trap sites, number of positive sites, number of negative sites and the total number of adults caught at each of the rural, urban or suburban land types. P-values are from Fisher Exact tests comparing the frequencies of positive/negative sites by land cover category, and total female catches by land cover category.

| *Cx. pipiens* |  |  |  |  |  |  |  |
| --- | --- | --- | --- | --- | --- | --- | --- |
| Land cover category | Number of trap sites | Number of sites positive | Number of sites negative | Test of association | Total females caught | Mean catch (SE) | Test of association |
| Rural | 139 | 54 (38.8%) | 85 (61.2%) | *P* < 0.001 | 910 (61.6%) | 16.6 (5) | *P* < 0.06 |
| Urban | 54 | 42 (77.8%) | 12 (22.2%) |  | 524 (35.5%) | 12.5 (2.8) |  |
| Suburban | 7 | 3 (42.9%) | 4 (57.1%) |  | 44 (3%) | 14.7 (5.8) |  |
| *Cx. molestus* |  |  |  |  |  |  |  |
| Rural | 139 | 2 (1.4%) | 137 (98.6%) | *P* = 0.28 | 2 (40%) | 1 (0) | *P* = 0.18 |
| Urban | 54 | 3 (5.6%) | 51 (94.4%) |  | 3 (60%) | 1 (0) |  |
| Suburban | 7 | 0 (0%) | 7 (100%) |  | 0 (0%) | NA |  |
| *Cx. pipiens/molestus* hybrids |  |  |  |  |  |  |  |
| Rural | 139 | 10 (7.2 %) | 129 (92.9%) | *P* = 0.64 | 17 (77.3%) | 1.7 (0.7) | *P* = 0.55 |
| Urban | 54 | 4 (7.4%) | 50 (92.6%) |  | 4 (18.2%) | 1 (0) |  |
| Suburban | 7 | 1 (14.3%) | 6 (85.7%) |  | 1 (4.5%) | 1 (0) |  |
| *Cx. torrentium* |  |  |  |  |  |  |  |
| Rural | 139 | 18 (12.9%) | 121 (87.1%) | *P* = 0.13 | 35 (92.1%) | 1.9 (0.3) | *P* = 0.01 |
| Urban | 54 | 2 (3.7%) | 52 (96.3%) |  | 3 (7.9%) | 1.5 (0.5) |  |
| Suburban | 7 | 0 (0%) | 7 (100%) |  | 0 (0%) | NA |  |

\*statistically significant

Individual catches of each species, by land cover category, are shown as boxplots in Figure 5. In the three land cover categories (rural, urban, suburban) 910 (61.6%), 524 (35.5%) and 44 (3%) *Cx. pipiens* were caught respectively (Table 2); there was a higher than expected number in urban sites although it was not significant (*P*=0.056). Mean catch sizes also did not differ significantly (Kruskal Wallace rank sum test, ns). These results suggest that whilst a higher proportion of urban sites caught *Cx. pipiens*, there is no effect of land cover on abundance, when the species is present.

**Fig. 5.**
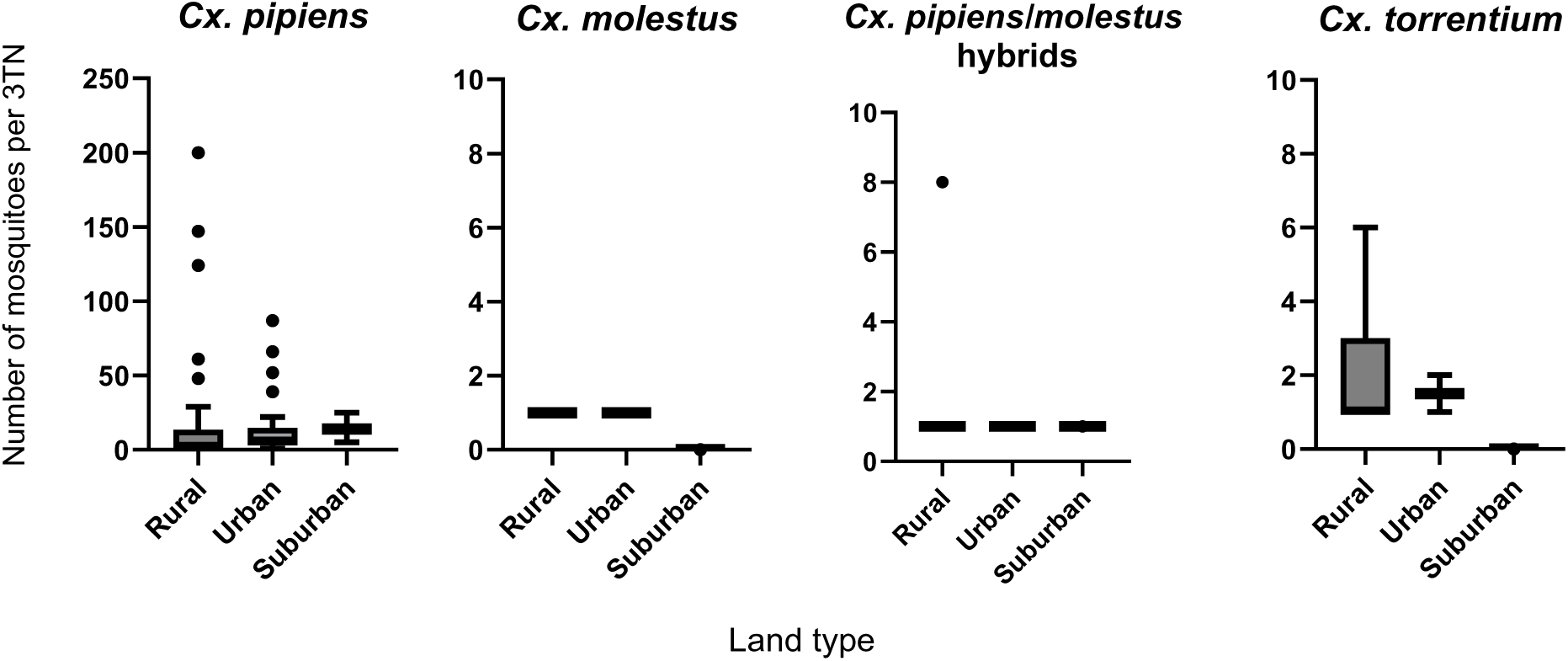
The number of *Cx. pipiens*, *Cx. molestus*, *Cx. pipiens/molestus* hybrids and *Cx. torrentium* found per 3TN at their rural, urban or suburban trap sites. Bars represent the mean number of mosquitoes found at rural, urban or suburban land types. Whiskers indicate interquartile ranges whilst dots represent individual data points that fall outside the IQR.

Although only 5 *Cx. molestus* were caught, 2 of them (40%) were from rural land types whilst 3 (60%) were from urban land types. This suggests no preference for land cover types (*P*=0.18).

We caught 17 (77.3%), 4 (18.2%) and 1 (4.5%) *Cx pipiens/molestus* hybrids at rural, urban and suburban sites respectively (*P*=0.55) indicating no preference towards land cover types.

We caught 35 (92.1%), 3 (7.9%) and 0 (0%) *Cx. torrentium* at rural, urban and suburban sites respectively (*P*=0.01) indicating a strong preference for rural environments. This effect is likely related to more rural sites being positive for *Cx. torrentium* rather than this species being more abundant in rural sites, when present; mean catches were slightly higher in urban sites although there were insufficient data from urban sites to draw reliable conclusions regarding mean catch sizes.

### Mosquito association to human populations and artificial structures

**Fig. 6.**
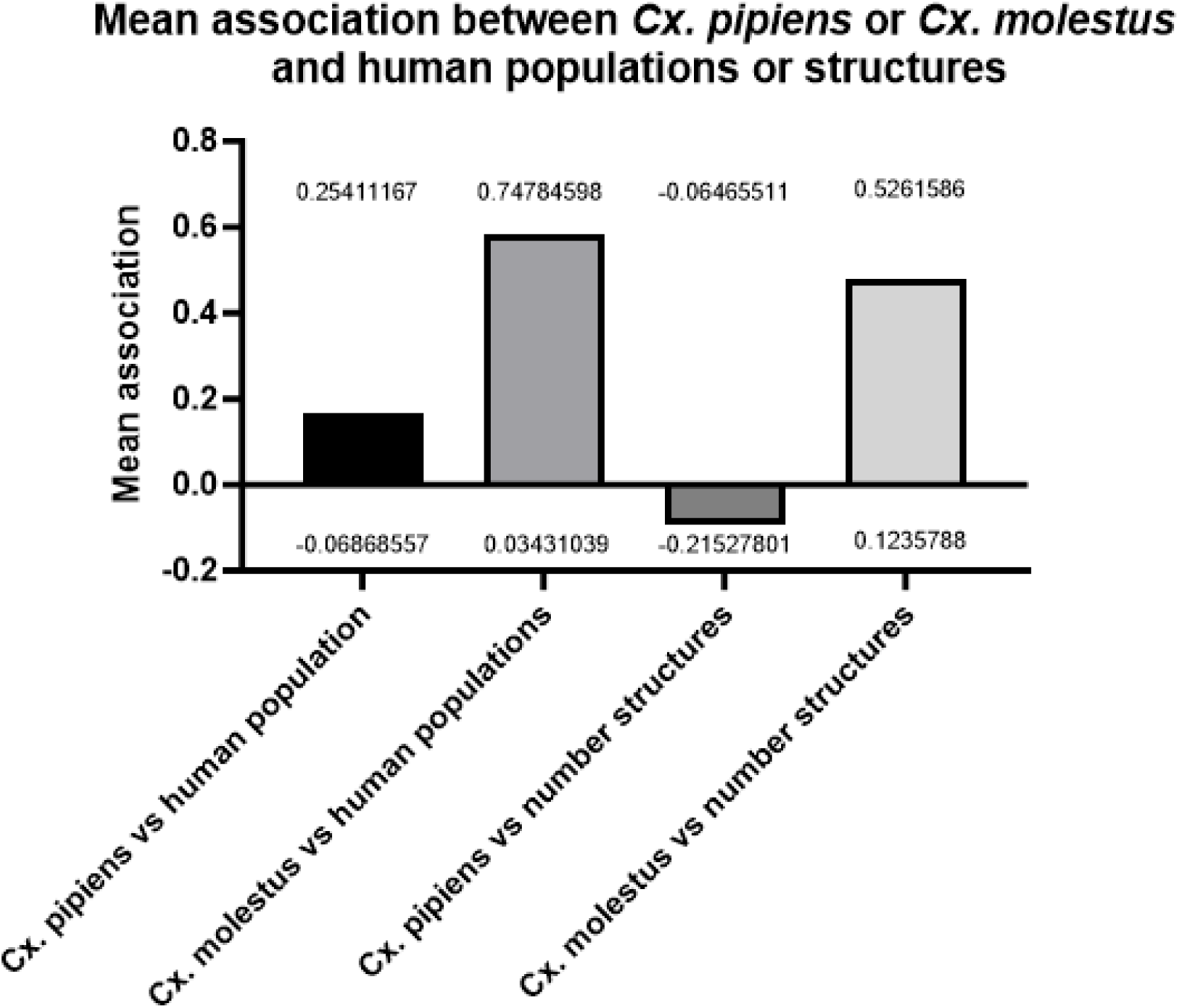
The values of association (measured as Pearson correlation within a Monte Carlo test) of *Cx. pipiens* and *Cx. molestus* to either human populations or number of structures (sum of the number of artificial structures, such as buildings, animal shelters, electricity infrastructures etc, within 500m from the trapping location). Bars represent the mean association between the two *Culex* mosquito biotypes, and human populations or artificial (man-made) structures. The upper and lower confidence intervals (UCI) and (LCI) are shown with the values printed above/below each bar. Number of structures refers to the total number of any artificial (man-made) structures within a 500m radius of the trap site.

There is a statistically significant and positive association between the presence of *Cx. molestus* and human populations (with an upper confidence interval of 0.74) and the presence of *Cx. molestus* and number of structures (Figure 5). There is a statistically significant negative association between *Cx. pipiens* and artificial structures (as identified by OpenStreetMap) (UCI −0.12). No statistically significant association between *Cx. pipiens* and human densities was found.

### Larval and pupal sampling

**Table 3.** The species identification of larvae collected from suitable sites (including water troughs/butts, buckets or tyres that can retain water and excluding rainwater puddles) within 25-100m of the traps. First instar larvae from each site were pooled to increase biological material in the DNA extraction (the 206 identified as *Cx. pipiens* complex included one pool and the 278 identified as *Cx. pipiens* included one pool). All larval data presented here came from the 100 trap sites in the south of England and Wales. P-values are from Fisher Exact tests comparing the frequencies of positive/negative sites by land cover category, and total larval catches by land cover category.

| Land type | Number of trap sites | Number of sites positive | Number of sites negative | Test of association | Total caught larvae | Mean catch (SE) | Test of association |
| --- | --- | --- | --- | --- | --- | --- | --- |
| *Cx. pipiens* |  |  |  |  |  |  |  |
| Rural | 71 | 7 (9.85%) | 64 (90.14%) | *P* = 0.46 | 27 (9.71%) | 3.86 (1.16) | *P* < 0.0001 |
| Urban | 23 | 4 (17.39%) | 19 (82.61%) |  | 99 (35.61%) | 24.75 (9.82) |  |
| Suburban | 6 | 1 (16.67%) | 5 (83.33%) |  | 152 (54.68%) | 152.00 (NA) |  |
| *Cx. pipiens/molestus* hybrids |  |  |  |  |  |  |  |
| Rural | 71 | 0(0%) | 71 (100%) | *P* = 0.03 | 0 (0%) | 0 | *P* < 0.04 |
| Urban | 23 | 1 (3.35%) | 22 (95.65%) |  | 1 (50%) | 1 |  |
| Suburban | 6 | 1 (16.67%) | 5 (83.33%) |  | 1 (50%) | 1 |  |
| *Cx. torrentium* |  |  |  |  |  |  |  |
| Rural | 71 | 2 (2.82%) | 69 (97.18%) | *P* = 1 | 10(90.91%) | 5 (0) | *P* = 0.6 |
| Urban | 23 | 1 (4.35%) | 22 (95.65%) |  | 1(9.09%) | 1 |  |
| Suburban | 6 | 0 (0%) | 6(100%) |  | 0 | 0 |  |

To ensure that the traps used in the study were not biased towards *Cx. pipiens*, given this biotype dominated the catches, we collected larvae and pupae at sites with suitable bodies of water within range of the traps. As with the traps, we saw a dominance of *Cx. pipiens* in the larvae/pupae. Equally, we identified *Cx. torrentium* larvae/pupae at three sites, one of which a site where adult *Cx. torrentium* were found. The hybrid larvae/pupae were not found at adult hybrid sites.

There was no evidence of an association between the presence of *Cx. pipiens* complex, or *Cx pipiens,* and land cover type (*P* = 0.46). However, the total number of larvae identified as *Cx pipiens complex*, or *Cx pipiens*, was significantly higher at suburban sites (*P* < 0.0001).

Only two *Cx. pipiens/molestus* hybrid larvae were found with none identified at rural sites, and one each at an urban and suburban. Given 71 sites were rural, this suggests a tendency to be present (*P* = 0.03) and more abundant (*P* = 0.04) at non-rural sites but catches were too small to have high confidence in this result.

Finally, there was no association between *Cx. torrentium* larval presence, or abundance, and land cover type although, as for hybrids, the small numbers found limit confidence in this result. The larval data from the south shows dominance of *Cx. pipiens* and suggests that our traps are not biased towards this biotype.

### Probability maps for Cx. pipiens and Cx. torrentium

**Fig. 8.**
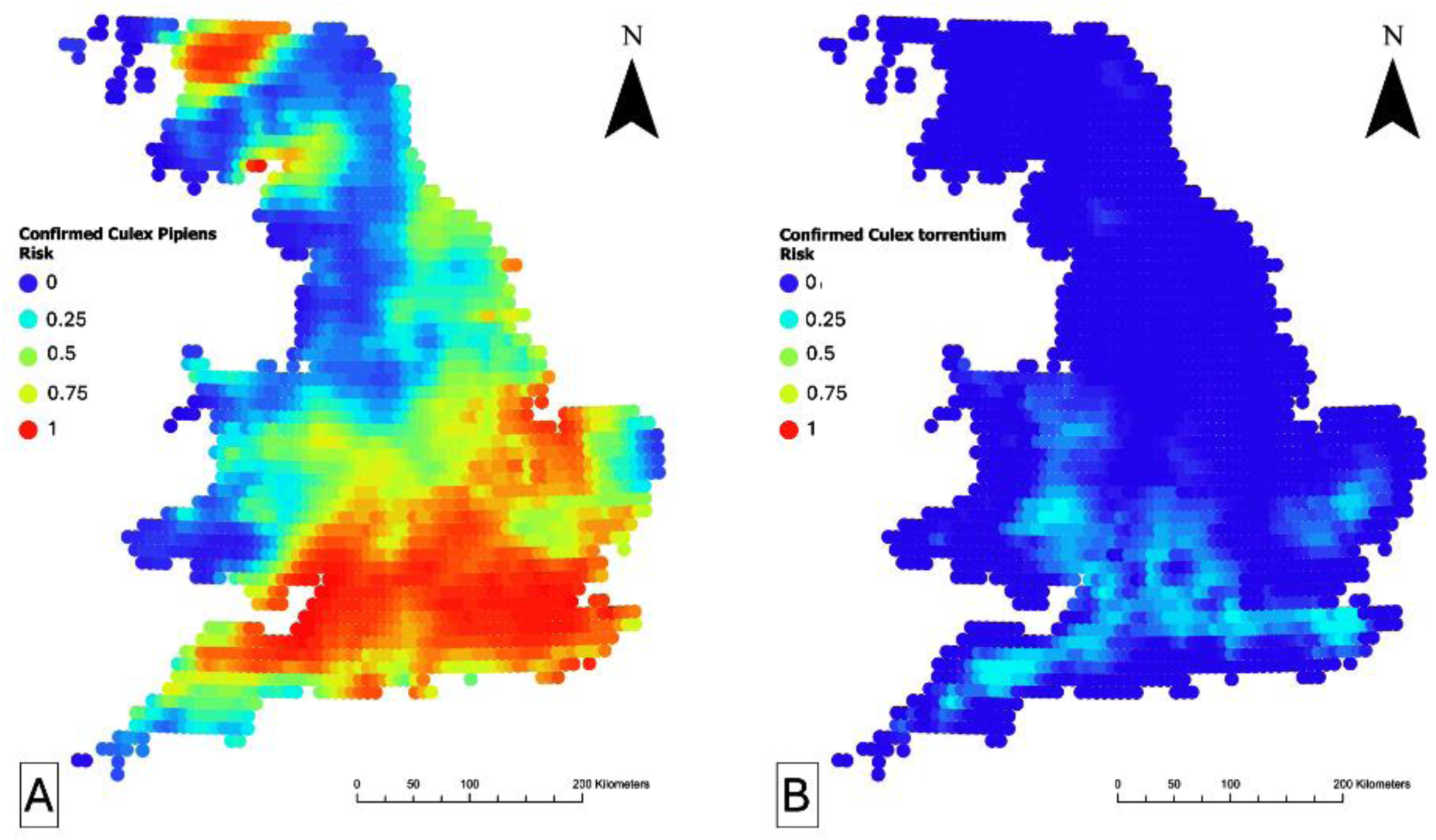
The probability of detecting *Cx. pipiens* and *Cx. torrentium* across England and Wales following the July 2023 field survey. A: the probability of detecting *Cx. pipiens,* B: the probability of detecting *Cx. torrentium.* Red indicates a probability of 1; blue indicates a probability of 0. *Cx. pipiens* represents *Cx. pipiens*, not the *Cx. pipiens* complex.

There is a non-zero probability of detecting *Cx. pipiens* in most of England and Wales apart from some coastal, and internal corridors, especially in the north. An exception is south Scotland and north Lakes, where interestingly the model predicted high detection risk. *Cx. torrentium* has a general lower risk of detection, compared to *Cx. pipiens* and mostly concentrated in the south-centre of England and Wales borders where we caught fewer *Cx. pipiens*.

## Discussion

To identify areas most at risk of outbreaks of WNV and other mosquito-borne diseases transmitted by *Culex* mosquitoes in the coming years, it is imperative we understand the distribution of the UKs competent mosquito vectors. Here, we present a robust dataset from the largest UK *Culex* mosquito spatial survey to date, conducted in July 2023.

### Cx. pipiens

The study identified a wide distribution of *Cx. pipiens* across England and Wales, with a higher density in the south of England. We found overwhelming predominance of *Cx. pipiens* amongst our confirmed biotypes, making up 95.8% of all species-confirmed individuals. This is consistent with other studies from northern and southern Europe. When looking at catch ratios of *Cx. pipiens* to *Cx. pipiens/molestus* hybrids, [65] found a decreasing gradient of *Cx. pipiens* from northern to southern latitudes across Europe, with 90% of the catch in Sweden *Cx. pipiens* compared to only 40% of catch in Italy. *Cx. pipiens* positive trap sites were evenly spread throughout the majority of England and Wales, though sparser in the north towards the England/Scotland border. There were clear regional differences across England and Wales regarding the number of *Cx. pipiens* caught, with higher numbers of *Cx. pipiens* on average at southern sites than northern sites, an important factor given the south of the UK has temperatures within the thermal limits for USUV transmission [28]. Despite the lower number of urban sites visited, a higher percentage of urban sites were positive for *Cx. pipiens*, whilst *Cx. pipiens* positive rural sites had a higher number on average of *Cx. pipiens*, though not significant. This may account for the lack of positive association to human populations or artificial structures seen with *Cx. pipiens*.

### Culex torrentium

*Cx. torrentium* is recognised as a primary enzootic vector for WNV, displaying the highest competence among the *Culex* species. When field caught mosquitoes in Germany were fed WNV [27], they observed transmission rates (the number of WNV RNA saliva positives as a proportion of the number of positive bodies) [27] of 90% and a transmission efficiency (the number of WNV RNA saliva positives as a proportion of individuals that blood-fed) of 24%. Additionally, [66] demonstrated *Cx. torrentium* can also serve as a competent vector for overwintering USUV. Knowledge of *Cx. torrentium* distribution in the UK is limited due to challenges in distinguishing it from the *Cx. pipiens* complex, though several recent studies have found the mosquito in the Midlands, North Yorkshire and London [67,28]. Our study is the first comprehensive survey of *Cx. torrentium* distribution across England and Wales. The prevalence and density of *Cx. torrentium* were much lower than *Cx. pipiens* (only 2.5% of the total species identified compared to 95.8% for *Cx. pipiens*) and its distribution was skewed towards south-centre of England and Wales borders. Notably, *Cx. torrentium* appears in several locations across Wales, areas where its presence was previously not studied, though it has been found in Anglesey previously [68]. To ensure the trap catch was not biased towards *Cx. pipiens*, larvae and pupae were collected at sites with suitable water bodies. The larval results agreed with the trap results.

### Culex modestus and invasive Aedes

*Cx. modestus* and *Cx. molestus* are two prominent bridge vectors for WNV in Europe [25,65,69]. Whilst no *Cx. modestus* was identified during this study, previous and more focused surveys have identified *Cx. modestus* across multiple sites at varying densities and populations have been shown to reach a peak during early August [45, 70]. *Culex modestus* has been recorded in Cambridgeshire, Dorset, Kent [45], Essex, Hampshire, Sussex, and Suffolk, and whilst this study did have sites within the known range of *Cx. modestus,* trap sites were not in the specific habitat known for the mosquito and as a result it was not recorded. This instead shows that these mosquitoes are not widely common in the rural and urban areas surveyed. It also showed the absence of other potential invasive species, such as *Aedes albopictus* and *Aedes japonicus*, both which are colonising mainland Europe. Though the importance of this is uncertain given the chance our trapping methods may have missed these.

### Culex molestus

Due to their human biting, *Cx. molestus* are capable of spreading WNV or USUV to humans if infected. Though *Cx. molestus* has been previously found in Neston, Cheshire [28], our study found *Cx. molestus* exclusively in the south of England. This is consistent with our understanding of *Cx. molestus* preferring warmer areas and previous findings in the UK and Europe [38,71]. However, given the low number of *Cx. molestus* found in the catches, and the greater number of mosquitoes found in the south of England and Wales, it is possible *Cx. molestus* are present but were not detected further north. Equally, overground populations of *Cx. molestus* have been reported in warmer areas of the UK and Europe. For instance, *Cx. molestus* overground populations have been found in London [76], and more *Cx. molestus* are found above ground in the south of Europe, in countries such as Italy [71]. Hence, they may be more likely to be detectable in above ground catches in the south of the UK than in the north.

A total of five *Cx. molestus* were detected during this study across five sites in the south of England (0.3% of the total identified catch). *Cx. molestus* was detected in Exeter, Devon at a trap site located by the edge of a park in a housing estate, near to a large area of allotments. *Cx. molestus* was also found in three sites in the southeast of England: on the eastern outskirts of Ipswich, Suffolk (housing estate with woodlands and river), in Southend-on-Sea, Essex, (burial ground in housing estate) and in Gravesend, Kent (housing estate near River Thames and sewage works). This finding of *Cx. molestus* adjacent to a sewage works in Kent is not unexpected, given the species association with sewage works in west (Mogden) and east (Beckton) London. With a positive association of *Cx. molestus* to both human populations and artificial structures, this corroborates that *Cx. molestus* can survive in artificial water containers around areas of housing, putting them in proximity to people. The final *Cx. molestus* site was in the mid- to southwest of England in rural Upton Scudamore, Wiltshire. This site is located next to a farm on the outskirts of a small village, with large open spaces of crop fields with no apparent large water bodies. *Cx. molestus* has previously been found on farms, though within the vicinity of Beckton sewage works in East London [38]. The presence of *Cx. molestus* on a rural farm confirms that this mosquito can survive in predominantly rural areas, potentially finding suitable habitats in nutrient rich farm associated water bodies. In contrast, the findings of *Cx. molestus* at Gravesend adjacent to a sewage works is not unexpected given the species association with sewage works in west and east London.

Multiple studies have shown differences in habitat for *Cx. pipiens* and *Cx. molestus*, with *Cx. molestus* identified as preferring underground or dark indoor habitats. Summarising results from several studies, [71] found that the divide between above-ground (*Cx. pipiens* habitats) and below-ground (*Cx. molestus* habitats) was more striking in northern latitudes and that this gradually decreases into mixed populations of *Cx. pipiens* and *Cx. molestus* at southern latitudes. It is possible the catches in this study were biased towards *Cx. pipiens* and *Cx. torrentium*, as no underground habitats were sampled therefore potentially missing *Cx. molestus* sites. However, our study does provide a useful indication of above ground species in both rural and urban areas.

Aside from the rural site in Wiltshire, several of the *Cx. molestus* sites are located within 5km of nearby wetlands where migratory birds spend the summer. As the ornithophilic *Cx. pipiens* is common across England and Wales, anywhere that *Cx. molestus* occurs in the presence of high densities of *Cx. pipiens* and infected birds, acts as a potential source for WNV transmission to humans. However, the low incidence of *Cx. molestus* and *Cx. pipiens/molestus* hybrids suggests that these locations may not be common, though potential foci for transmission by *Cx. molestus* do exist.

### Cx. pipiens/molestus hybrids

In our study, there were low numbers of *Cx. pipiens/molestus* hybrids (n=22), making up 1.4% of the total confirmed catch, across fifteen trap sites. All hybrid positive sites had only one identified hybrid, other than the site in Tiverton, Devon, which had *eight*. As with *Cx. molestus*, the hybrid sites were almost exclusively in the south of England and one site in south Wales (Cardiff). Three of the hybrid sites were sites where we also found *Cx. molestus* (Exeter, Ipswich and Upton Scudamore), which suggests that interbreeding between *Cx. pipiens* and *Cx. molestus* is occurring at these sites. Including the site shared with *Cx. molestus* at Upton Scudamore, five (out of 15) hybrid sites were outside farms in rural areas (Holnest [Dorset], Dengie [Essex], Tiverton [Devon] and East Kennet [Wiltshire]). This suggests that *Cx. pipiens/molestus* hybrids may breed and survive in the nutrient rich water sources found at farms, similar to the *Cx. pipiens* hybrids found around the farms and botanical gardens of Neston, Cheshire [28]. Five of the sites are within or near cities with sites next to rivers: Bristol next to the River Avon, Cardiff in Bute Park next to the River Taff; Upper Stoke (near Gravesend) in between the estuaries for the River Thames and River Medway; and Padstow (Cornwall) at the River Camel. These provide sites near water where hybrids are in direct contact with human populations. The final two sites include Toys Hill (Kent), a large, wooded area and Thaxted (Essex), on the edge of the two near a large area of crop fields.

It is not certain whether *Cx. molestus* were missed at the sites where *Cx. pipiens*/molestus hybrids (if hybrids are offspring from *Cx. pipiens* x *Cx. molestus* crosses) were found, or whether hybrids generate further populations of hybrids without the need for *Cx. pipiens* or *Cx. molestus* presence in nature. It is worth noting our lab maintains a colony of hybrids, suggesting they can produce hybrid populations. If the latter is true, this suggests that the hybrids can survive in a wider range of environments suited to *Cx. pipiens*, whilst *Cx. molestus* may still be limited in its range. It is worth noting that [59] state the assay used to detect hybrids will identify first generation hybrids, whilst backcrossing of the biotypes will result in recombination of the target site, presumably leading to further *Cx. pipiens/molestus* hybrid crosses that cannot be identified using our methods. Regardless, all hybrids in this study were found in the south of England and Wales, suggesting temperature could still be a limiting factor in their spread.

WNV outbreaks occur predominantly in south and central Europe despite competent mosquito and bird species present in northern Europe, though virus outbreaks have occurred in countries such as Germany in recent years. Multiple studies alluded to temperature as the reason for this disparity rather than regional differences in the species of *Culex* mosquito [65,72,73,69,27]. Evidence suggests that competent vector species of mosquito are present in northern Europe however at the colder temperatures the extrinsic incubation period of the viruses will be longer, potentially too long for sustained transmission. Temperatures above 21 degrees Celsius support WNV transmission and could lead to short summertime outbreaks in northern Europe. When modelling temperatures deemed suitable for WNV establishment across countries in Europe, almost all countries had at least one month of WNV permissive temperature in 2020, with the south of the UK having around two months [74]. With global warming increasing temperatures, the conditions are becoming more supportive for WNV transmission in the UK. With USUV already endemic in the south of England, UK species can act as competent enzootic vectors for the virus. To date, no published studies have investigated the vector competence of UK *Culex* mosquitoes for WNV virus, though *Cx. pipiens* has been shown to be competent for Japanese encephalitis virus, a flavivirus closely related to WNV [75]. Understanding their distribution in the UK will be key to predicting the locations of future outbreaks once the climate becomes suitable for WNV and further USUV spread. While temperature plays a key role in competence, the proximity to migratory birds and human populations along with the number of mosquitoes is also important [28] provides UK *Cx. molestus* mosquito vector competence data for USUV which can be integrated into a modelling approach alongside our data to provide a risk map for USUV. Furthermore, through risk modelling this data can be used to predict areas most at risk of WNV emergence, establishment and spread and highlight areas where continued surveillance is required.

## Conclusion

This study is the first comprehensive field survey of the *Cx. pipiens* complex and *Cx. torrentium* across England and Wales. *Cx. pipiens*, a prime enzootic vector for WNV and USUV, is widespread across England and Wales, and is the dominant *Culex* species in both adult catch and larval sampling. Although *Cx. torrentium* was detected, it only accounted for a small proportion of the trapped *Culex* mosquitoes. We identified several locations with *Cx. molestus* that warrant further study, and the presence of hybrids of both *Cx. pipiens* and *Cx. molestus* in the south of the UK. This paper provides data on mosquito distribution that can be used to support USUV and WNV risk modelling to inform future research and preparedness on the threat of Culex-borne arboviruses in the UK.

## Data Availability

All data produced in the present work are contained in the manuscript, or are avalaible upon request to the authors

## Supplementary information

**Table S1.** A list of the non-*Culex* mosquito species identified morphologically during the 2023 field survey.

| **Mosquito Species** |
| --- |
| *Aedes cantans/annulipes* |
| *Aedes cinereus* |
| *Aedes detritus* |
| *Anopheles atroparvus* |
| *Anopheles claviger* |
| *Anopheles maculipennis s.l.* |
| *Anopheles plumbeus* |
| *Coquillettidia richiardii* |
| *Culiseta annulata* |
| *Culiseta litorea* |
| *Culiseta morsitans* |
| *Unidentifiable* |

**S2.**
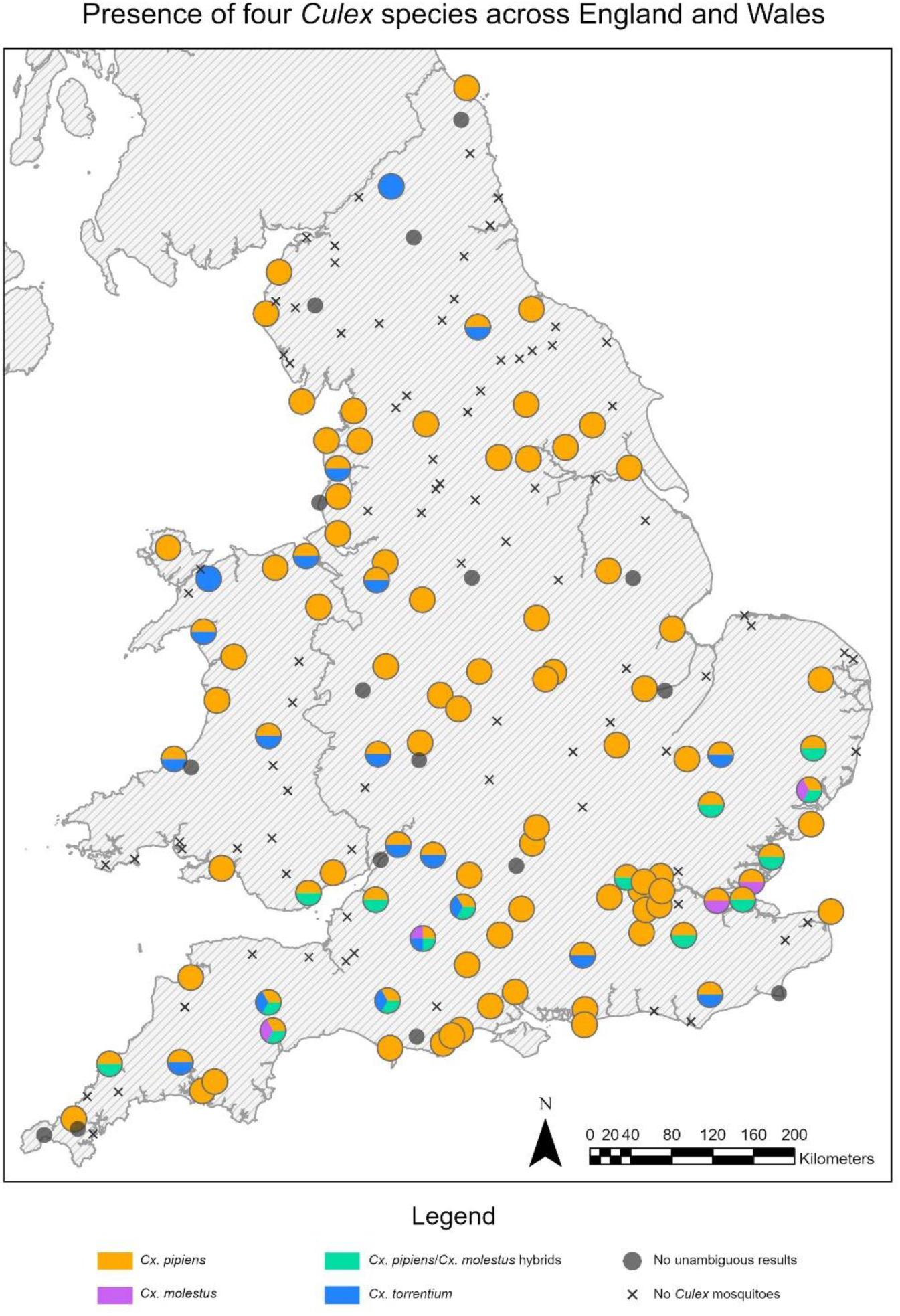
A map showing an equal divide of each of the four *Culex* species found at each site, i.e. pure orange squares represent sites where only *Cx. pipiens* was found, whilst orange, pink, green and blue circles represent sites where all four of the *Culex* species were found.

## Availability of data and materials

All data are included in the manuscript and supplement

## Author’s contributions

Conceptualization: LS, JM, MB, MSCB. Methodology: EW, LS, JM, MB, MSCB. Experimentation: RW, EW, AV, JTH, AD, AS, AJA, CJJ, JM, KB, EA, SS, SG, SMB. Sample preparation: RW, EW, AV, JTH, KS, AHR. Project administration: EW, JM, MB, MSCB. Data curation: RW, EW, AHR. Visualization: RW, EW, AHR, LS. Writing—review and editing: All authors.

## Ethics approval and consent to participate

No ethical consideration required

## Consent for publication

Not applicable.

